# Revealing public opinion towards COVID-19 vaccines with Twitter Data in the United States: a spatiotemporal perspective

**DOI:** 10.1101/2021.06.02.21258233

**Authors:** Tao Hu, Siqin Wang, Wei Luo, Mengxi Zhang, Xiao Huang, Yingwei Yan, Regina Liu, Kelly Ly, Viraj Kacker, Bing She, Zhenlong Li

## Abstract

**Background:** The COVID-19 pandemic has imposed a large, initially uncontrollable, public health crisis both in the US and across the world, with experts looking to vaccines as the ultimate mechanism of defense. The development and deployment of COVID-19 vaccines have been rapidly advancing via global efforts. Hence, it is crucial for governments, public health officials, and policy makers to understand public attitudes and opinions towards vaccines, such that effective interventions and educational campaigns can be designed to promote vaccine acceptance.

**Objective:** The aim of this study is to investigate public opinion and perception on COVID-19 vaccines by investigating the spatiotemporal trends of their sentiment and emotion towards vaccines, as well as how such trends relate to popular topics on Twitter in the US.

**Methods:** We collected over 300,000 geotagged tweets in the US from March 1, 2020 to February 28, 2021. We examined the spatiotemporal patterns of public sentiment and emotion over time at both national and state scales and identified three phases along the pandemic timeline with the significant changes of public sentiment and emotion, further linking to eleven key events and major topics as the potential drivers to induce such changes via cloud mapping of keywords and topic modelling.

**Results:** An increasing trend of positive sentiment in parallel with the decrease of negative sentiment are generally observed in most states, reflecting the rising confidence and anticipation of the public towards vaccines. The overall tendency of the eight types of emotion implies the trustiness and anticipation of the public to vaccination, accompanied by the mixture of fear, sadness and anger. Critical social/international events and/or the announcements of political leaders and authorities may have potential impacts on the public opinion on vaccines. These factors, along with important topics and manual reading of popular posts on eleven key events, help identify underlying themes and validate insights from the analysis.

**Conclusions:** The analyses of near real-time social media big data benefit public health authorities by enabling them to monitor public attitudes and opinions towards vaccine-related information in a geo-aware manner, address the concerns of vaccine skeptics and promote the confidence of individuals within a certain region or community, towards vaccines.

## 1. Introduction

As of May 21, 2021, the COVID-19 pandemic has led to more than 160 million confirmed cases and more than three million deaths worldwide [1]. SARS-CoV-2 has been out of control in many parts of the world due to its highly contagious nature, diverse variants, and the mass public’s inconsistent adherence to effective public health measures, such as wearing masks and maintaining social distancing [2]. Meanwhile, the emergence of asymptomatic cases (which are difficult to detect) has become more frequent potentially leading to a substantial accumulation in the number of infections over time [3]. As such, the only hope of controlling the viral spread is the wide availability and public acceptance of safe and effective vaccines [4].

Since January 2020, scientists and medical experts around the world have been developing and testing COVID-19 vaccines; 16 vaccines have been approved for emergency use around the world by far, but the progress of vaccination has been subject to hesitancy, distrust and debate. Vaccine hesitancy has been identified by the World Health Organization (WHO) as one of the top ten global health threats in 2019 [5], and such hesitancy along with the misinformation of vaccination, have presented substantial obstacles towards achieving the sufficient coverage of vaccinated populations to realize community immunity in many countries [6,7].

Therefore, it is crucial for governments, public health officials, and policy makers to understand the potential drivers that affect public opinion towards COVID-19 vaccines [8]. A number of campaigns against the anti-vaccination activists have been made through multiple channels since January 2020. Notably, the accelerated pace of vaccine development has further heightened public anxieties and could compromise the public’s acceptance of the vaccine [9]. However, this acceptance varies across geographic contexts and the pandemic timeline. As governments put more effort into developing strategies for promoting vaccine acceptance and uptake, the key questions regarding vaccination willingness persist — what are the opinions and perceptions of the public to COVID-19 vaccines and what are the potential drivers that affect such opinions?

In COVID-19 vaccine related-studies, using big data from social media platforms such as Twitter has been an emerging trend in recent scholarship. Geotagged Tweets (hereinafter termed as geotweet) provide a rich volume of near real-time and cost-effective contents including news, events, public comments, and the locational information of Twitter users. Through sentiment analysis and topic modelling methods that have been widely used in existing studies, qualitative tweet contents can be retrieved to reflect public opinions and attitudes towards COVID-19 vaccines. Additionally, user locational information enables researchers to investigate the spatiotemporal patterns of the public’s opinions and attitudes. In general, existing studies have investigated people’s reactions towards COVID-19 vaccines, with a heavy focus on the US [10-15] and also extends to other countries in the world, including China [16], South Africa [17], Australia [18], United Kingdom [19-20], Canada [21, 11], Africa [22] and to a global scale [23]. However, the study period of these works is relatively limited to or predominantly focused on the early stage of the pandemic or up to the end of 2020, without covering early 2021, the period of mass systemic vaccine implementation in many countries. Furthermore, although sentiment analysis and topic modeling have been broadly applied, what remains less explored are the potential drivers that induce the change in public sentiment and opinion on vaccines, such as important events and the announcement of political leaders (e.g., the propaganda of vaccine success or vaccine conspiracy). There is a pressing need to investigate public opinion towards vaccines across a longer timeline and explore the potential drivers that influence the change of such opinion over time.

To address these knowledge gaps, this study aims to analyze the spatiotemporal patterns of the public sentiment and emotion and explore the keywords and major topics of tweets regarding COVID-19 vaccines among Twitter users in order to unveil the potential drivers relevant to the change of sentiment and emotion over time. Drawing on more than 300,000 geotweet from March 1, 2020 to February 28, 2021 in the US, we employed sentiment analysis and emotion analysis at the national and state level and identified three phases along the pandemic timeline that display significant changes in public sentiment and emotion, while linking to eleven key events and major topics as the potential drivers that induce such changes via cloud mapping of keywords and topic modelling. Findings from this study could help governments, policy makers, and public health officials understand public motivation and hesitance in order to design potential interventions to promote vaccination.

## 2. Data and Methodology

### 2.1 Data

We collected geotweet from March 1, 2020, to February 28, 2021, using the Twitter streaming Application Programming Interface (API). Geotweet provide the location information of the user-defined places when users post tweets or their longitude and latitude, if they activate the Global Positioning System (GPS) function in Twitter. The keyword “vaccin*” was used to search via the Twitter API, generating a total of 308,755 geotweet relevant to vaccines. In the results, 1.43% (44,118) of geotweet’ geographic locations are at a state-level (i.e., MA, USA), and others are geocoded at a city-level (i.e., Cambridge, MA) or a finer geographical level (i.e., Uptown Coffee, Oxford, MS). We then conducted a series of data preprocessing to the geotweet’ contents. First, we generalized the variations of COVID-related terms to “COVID-19”, including “corona”, “covid”, “covid19” and “coronavirus”; second, we removed the unrelated website links from the searching results, including the links starting from the fragment of “https”; third, we removed the punctuations (e.g., a period, question mark, comma, colon, and ellipsis) and other key symbols (e.g., a bracket, single and double quote) and converted the capital into lower case; fourth, we removed the inflectional endings (e.g., “ly”) and returned the root or dictionary form of a word by employing the function of *word lemmatization* provided in the Python package Natural Language Toolkit 3.6.2 [24].

### 2.2 Methodology

We conducted four sets of analyses to explore the spatiotemporal patterns of the public sentiment and emotion towards COVID-19 vaccines, including sentiment analysis, emotion analysis, topic modelling, and word cloud mapping. For the sentiment analysis, we applied Valence Aware Dictionary for Sentiment Reasoning (VADER), a well-known rule-based model, to estimate sentiment compound scores [25]. The sentiment compound score is computed by summing the score of each word in the lexicon, adjusted according to the rules, which embody grammatical and syntactical conventions for expressing and emphasizing sentiment intensity. Then the score is normalized to be between -1 (most extreme negative) and +1 (most extreme positive). To reclassify sentences to positive, neutral, or negative sentiment, threshold values are set as follows: a tweet with a compound score larger than 0.05 is classified as positive sentiment; a tweet with a compound score smaller than -0.05 is classified as negative sentiment; otherwise, it is classified as neutral sentiment [25]. We then cross-tabulated the three types of sentiment on daily and weekly basis along with the number of geotweet, and generated line graphs at the national level and in the top 10 states with the largest number of geotweet.

Differing from sentiment analysis, which detects positive, neutral, or negative feelings from tweet contents, the emotion analysis aims to recognize the types of feelings more specifically through the content expression, such as anger, fear, and happiness. The emotion analysis of this study was performed based on the National Research Council Canada Lexicon (NRCLex) [26]. NRCLex examines four pairs of primary bipolar emotions: joy (feeling happy) versus sadness (feeling sad); anger (feeling angry) versus fear (feeling of being afraid); trust (stronger admiration and weaker acceptance) versus disgust (feeling something is wrong or nasty); and surprise (being unprepared for something) versus anticipation (looking forward positively to something). We then examined the temporal patterns of these eight types of emotion at both national and state levels.

We identified 11 key dates (Figure 1) as the turning points of sentiment scores or with the significant change of the number of geotweet. In order to investigate the potential drivers of such changes, we applied Latent Dirichlet Allocation (LDA) model [27] to detect topics based on Twitter content. The LDA model generates automatic summaries of topics in terms of a discrete probability distribution over words for each topic, and further infers per-document discrete distributions over topics [28]. Each topic is treated as a cluster and each document will be assigned to a cluster which represents its dominant topic. LDA is an unsupervised algorithm [29], meaning that users do not know prior to running the model how many topics exist in the corpus, thus the number of topics are predefined by users. To estimate the optimal number of topics, we used the Python package [30] and pyLDAvis [31] to compare the results with topic numbers from 3 to 10, and found that the smallest overlap among topics occurs when the topic number is 3.

**Figure 1.**
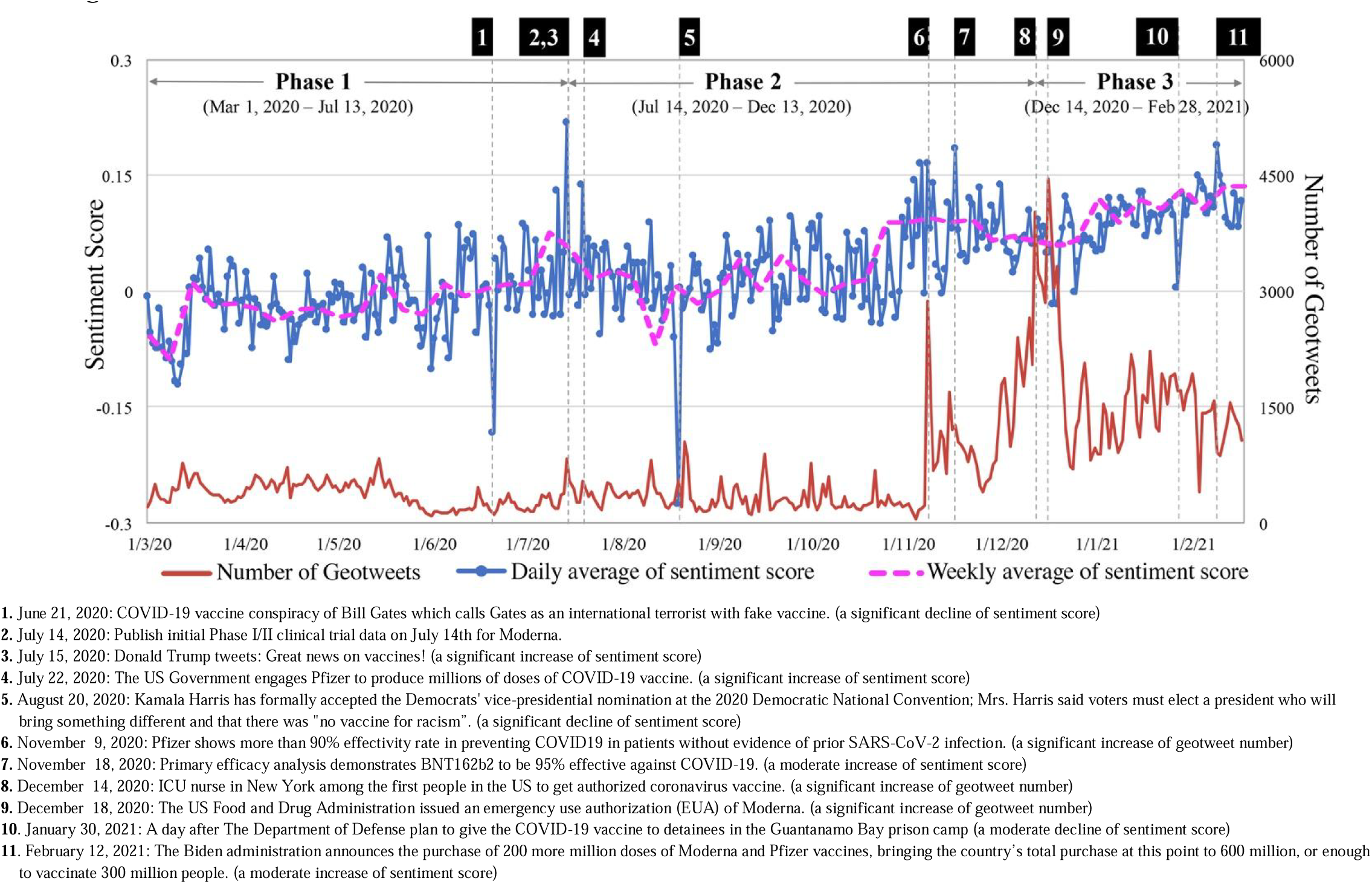
Sentiment scores and the number of geotweet over the entire study timeline with key dates

We further categorized the study period into three phases based on two iconic events: the results of Phase 1 clinical trials of Moderna were published in the *The New England Journal of Medicine* on July 14, 2020 [32] and the first COVID-19 vaccine shots were given in the US on December 14, 2020 [33]. Phase 1 from March 1, 2020 to July 13, 2020 is the stage in which the public was waiting for official announcements regarding the effectiveness of COVID-19 vaccines; Phase 2 ranges from July 14, 2020 to December 13, 2020 when positive news of COVID-19 vaccine development started coming; Phase 3 starts from December 14, 2020 when the first vaccine shots were given in the US. We then aggregated sentiment scores at the state level and analyzed the changes in sentiment over the three phases in the top 10 states. Finally, we produced word cloud maps over three pre-defined phases based on the frequency of keywords appearing in Tweets contents, with the size of a key word reflecting its frequency and popularity.

## 4. Results

### 4.1 Sentiment analysis and topic modeling

Figure 1 shows the overall trends of the weekly sentiment scores, unveiling the increased positive attitude towards COVID-19 vaccines within the study period. We then identified 11 key dates for when the sentiment scores or the number of geotweet changed dramatically. Topic modeling summarizes the three most popular topics discussed on each key date. A total of 33 topics on the 11 key dates are summarized and presented in Figure 2. Within each topic graph, the Y-axis indicates the top nine keywords associated with that topic and X-axis shows the frequency of each keyword, revealing to what extent a certain keyword contributes to that topic. Based on the top nine most relevant keywords to each topic, we generalized and presented a name of each topic at the bottom of each graph in Figure 2. We then identified key events of the selected dates through analyzing these topics.

**Figure 2.**
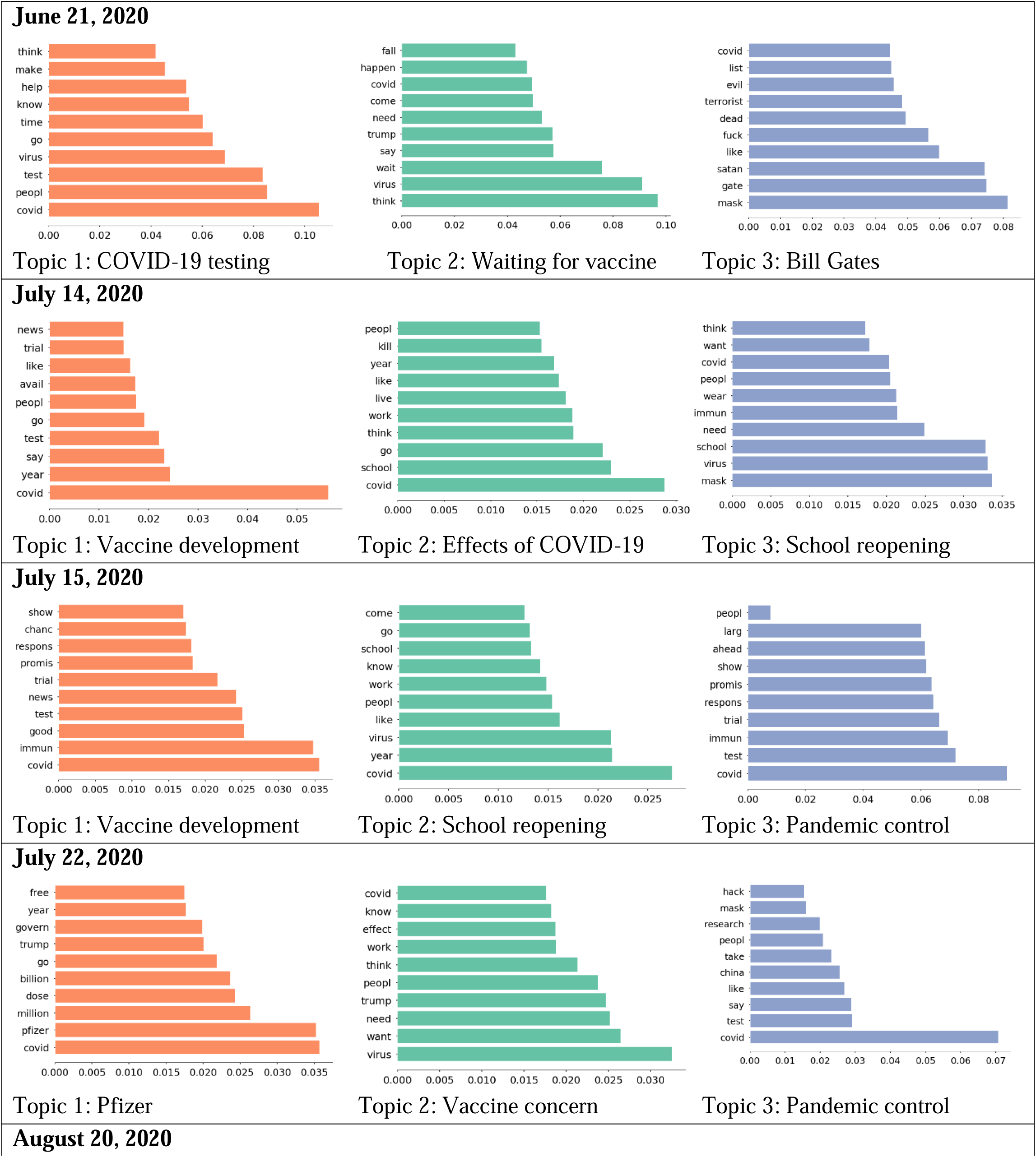
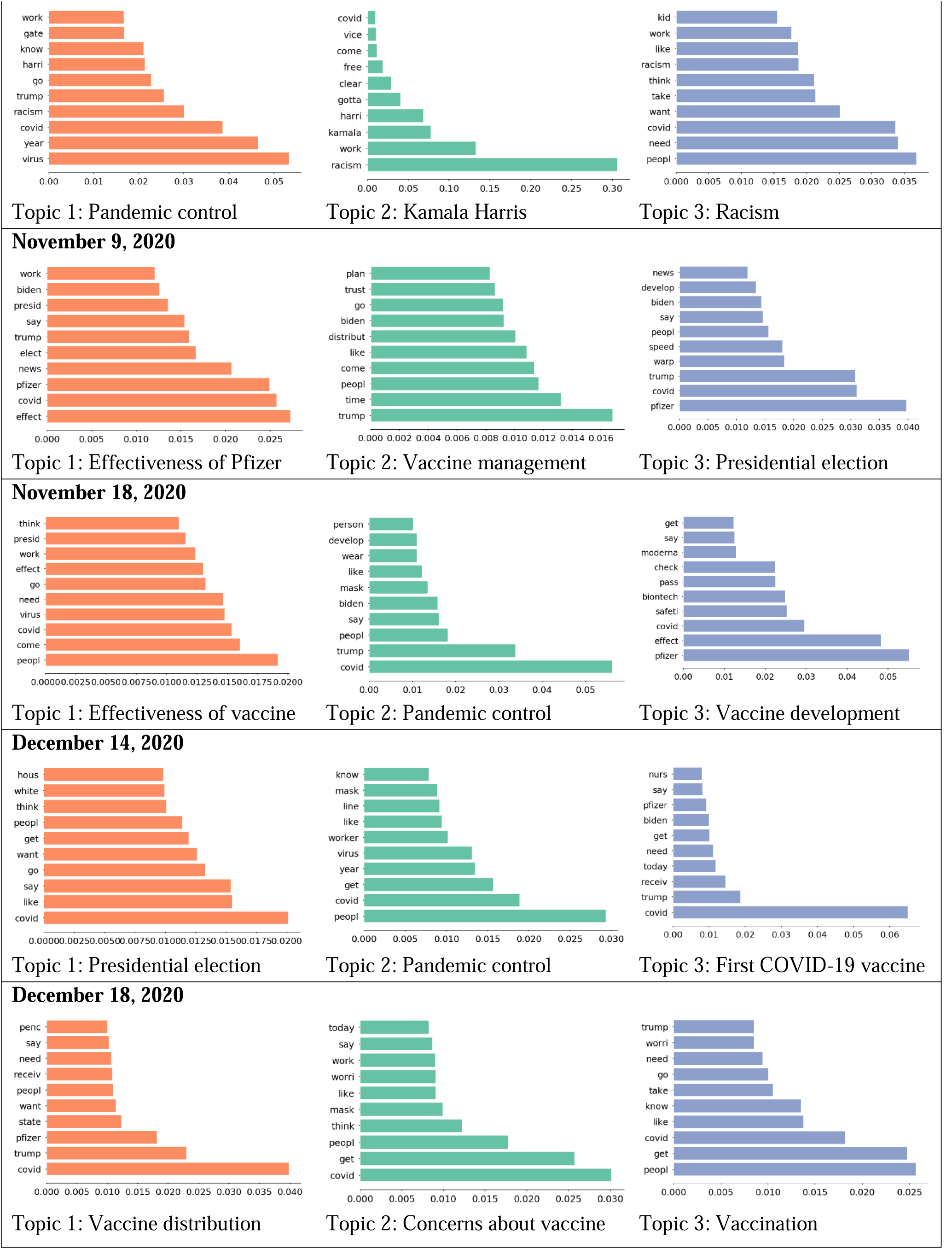
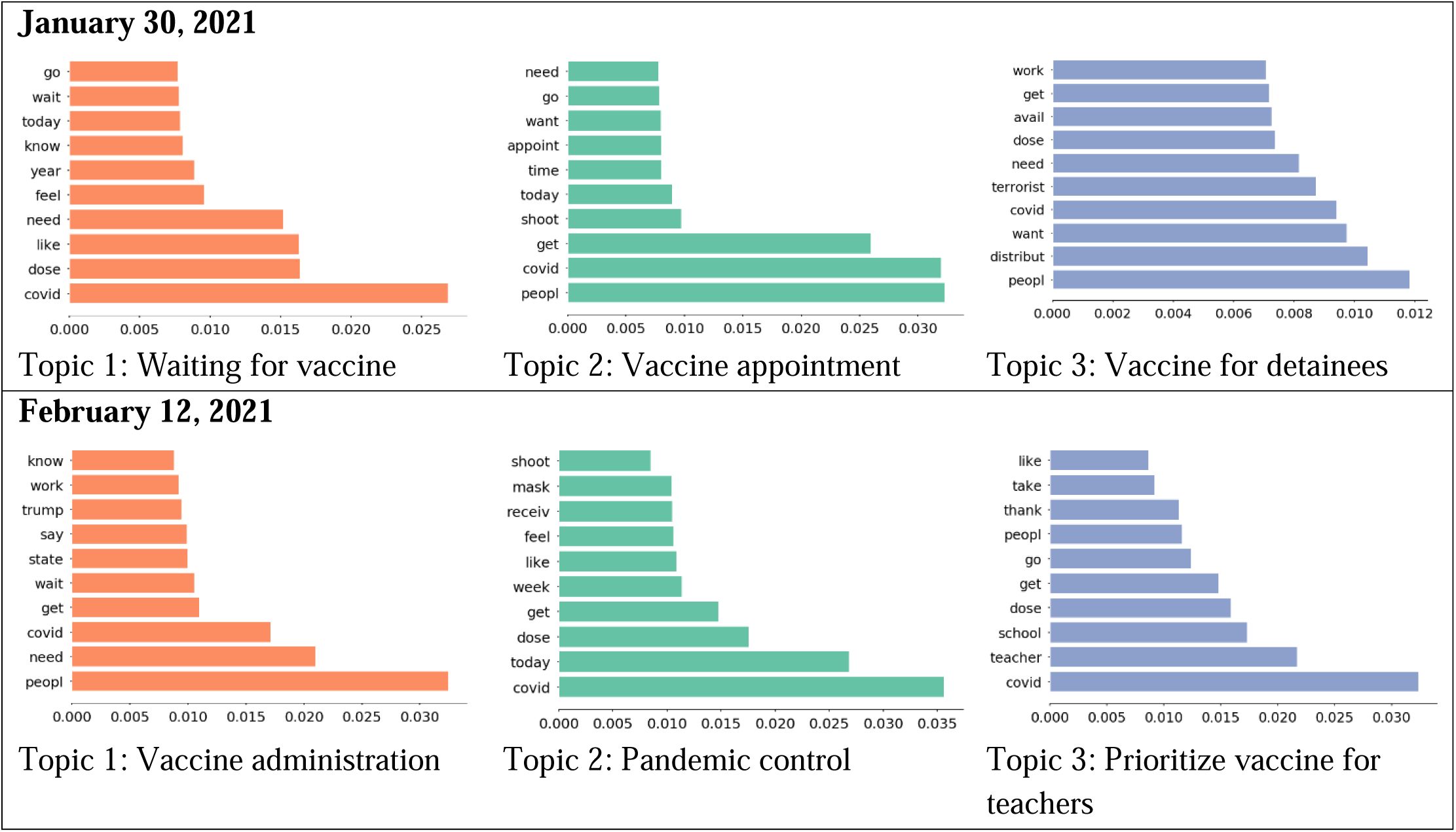
Top 3 topics discussed on eleven key dates Note: Within each figure, X axis indicates weight of each word; Y axis indicates top 10 words in the topic.

In Phase 1, changes in the sentiment score have been relatively stable, except for a sharp drop on June 21, 2020. This drop could have resulted from the misinformation and conspiracy theories related to Bill Gates. Vaccine-adverse conspiracy related to Gates claimed that the pandemic is a cover for his plan to implant trackable microchips made by Microsoft [34]. Topic modeling suggests that Gates was referred to as “satan”, “terrorist”, and “evil” on that day (Figure 2).

In Phase 2, the first stimuli was observed on July 14, 2020 when the results of Phase 1 clinical trials of Moderna were published [32], but we did not observe a dramatic change in sentiment score until July 15, 2020, when Donald Trump tweeted “Great News on Vaccines!” [32]. Topic modeling suggests that keywords related to “good”, “trial”, “promis”, and “test” were widely discussed on July 15, 2020 (Figure 2). Speculation suggests that comments from public figures on vaccination could trigger bigger changes in public sentiment compared with key events of the development of COVID-19 vaccines.

Another significant increase in sentiment score was observed on July 22, 2020, when the engagement of Pfizer with the US government accelerated the production and delivery of 100 million doses of COVID-19 vaccines [35]. The keywords “pfizer”, “govern”, and “million” were widely discussed and identified through topic modeling (Figure 2). On August 20, 2020, sentiment score dropped dramatically after Kamala Harris formally accepted the Democrats’ vice-presidential nomination at the 2020 Democratic National Convention, with the advocacy of “There is no vaccine for racism” naming victims like George Floyd and Breonna Taylor [36]. Of the keywords, “racism” and “kamala” were observed through topic modeling. Another increase in sentiment score appeared on November 9, 2020, when Pfizer announced that its vaccine is 90% effective (Figure 2) [37]. On the same day, Trump tweeted “STOCK MARKET UP BIG, VACCINE COMING SOON. REPORT 90% EFFECTIVE. SUCH GREAT NEWS!”. In the midst of positive news from Pfizer, people questioned whether Pfizer purposefully released study results after Election Day, though Pfizer’s CEO claimed that the releasing time has nothing to do with politics [38]. The widely discussed keywords include “trump”, “pfizer”, and “elect” on that day (Figure 2).

In Phase 3, an increased sentiment score was observed when an Intensive Care Unit (ICU) nurse received the first COVID-19 vaccine in New York on December 14, 2020. On the same day, the Electoral College voted to cement Biden’s victory over Trump. Discussion regarding COVID-19 vaccines (“pfizer”, “nurs”, “receive”) quickly raised on twitter, while other related discussions regarding mask wearing (“wear” and “mask”) and the presidential election (“house”, “trump”, “biden”) remained popular (Figure 2). By December 18, 2020, both Pfizer and Moderna were authorized for emergency use by The US Food and Drug Administration [39] and the sentiment score remained high. Trump tweeted “Moderna vaccine overwhelmingly approved. Distribution to start immediately,” Additionally, the fact that the former Vice President Pence and second lady Karen Pence received COVID-19 vaccine [40] was widely discussed (“penc” and “receiv”). Expectations for the COVID-19 vaccines were also discussed (“need” and “want”) (Figure 2). On January 30, 2021, The Department of Defense paused a plan to give COVID-19 vaccines to detainees in the Guantanamo Bay prison camp [41], which raised queries of COVID-19 vaccine delivery, leading to a moderate decrease in the sentiment score. Keywords were observed, including “terrorist” and “distribut” through topic modeling (Figure 2). On February 12, 2021, an increased sentiment score was observed after the Biden administration announced the purchase of 200 million COVID-19 vaccine doses from Pfizer and Moderna [42]. Discussion surrounding the administration of COVID-19 vaccines was initiated extensively (“wait”, “get”, “need”) (Figure 2). Topic modeling also suggests that teachers complained in the states in which they were not prioritized for vaccination, despite CDC’s recommendation (“teacher”, “school” and “get”).

We then broke down the sentiment scores by state in tandem along with the pandemic timeline. We presented the results in the top 10 states with the largest number of geotweet (Figure 3), including California, New York, Texas, Florida, Illinois, Ohio, North Carolina, Pennsylvania, Georgia, and Virginia. The temporal patterns of sentiment scores vary across states, with more obvious fluctuations before November 2020 in Illinois, Ohio, North Carolina, Georgia, Pennsylvania, and Virginia. A number of significant decreases of sentiment scores were observed in June 2020 in Illinois, North Carolina, Ohio, Pennsylvania, Georgia, and Virginia, in line with the tendency of sentiment drops at the national level. The states with relatively larger numbers of geotweet (i.e., California, New York, Texas, and Florida) are more stable in the temporal trends of sentiment scores, compared to the states with relatively smaller numbers of geotweet (e.g., Ohio, North Carolina, Pennsylvania, Georgia, and Virginia).

**Figure 3.**
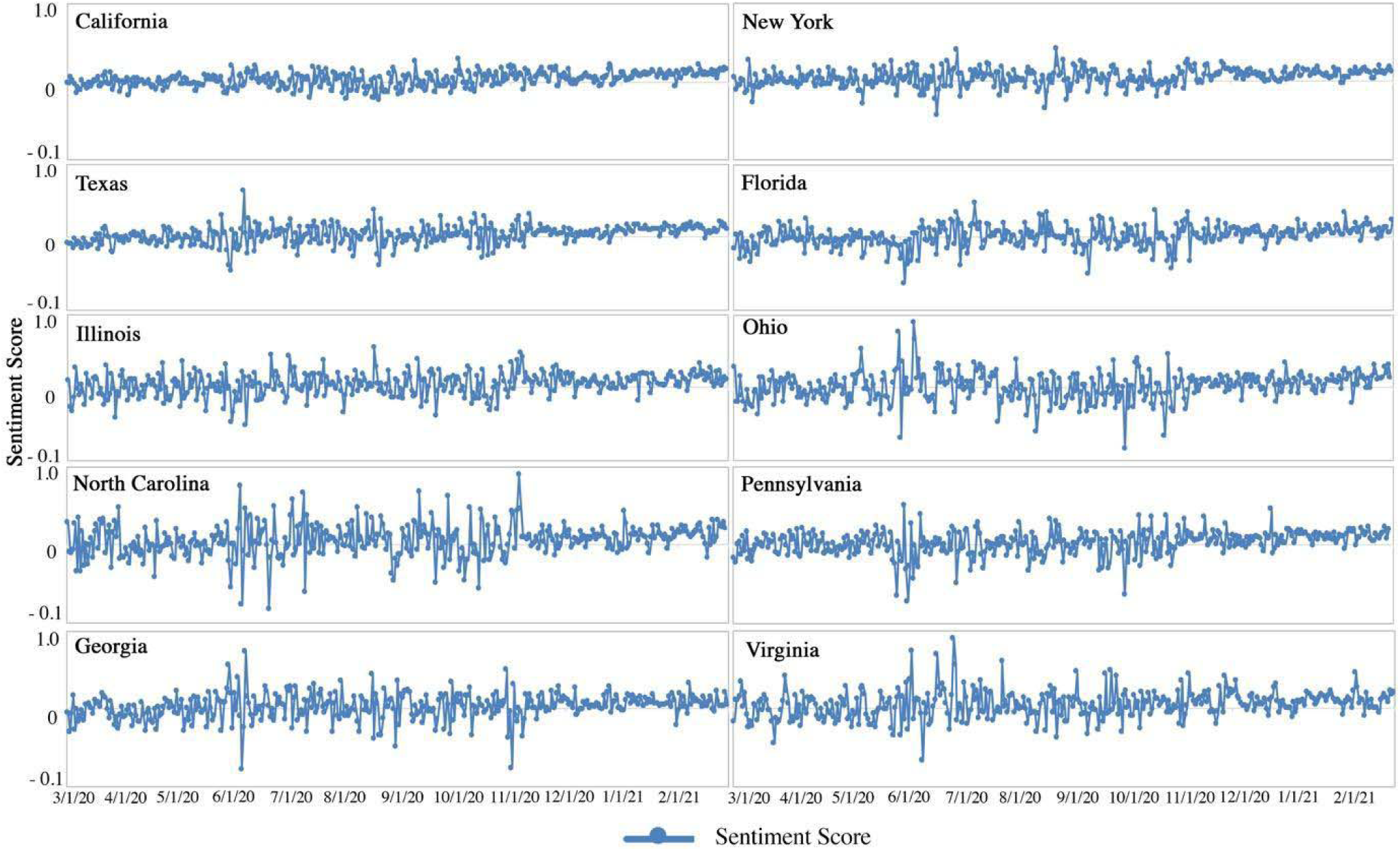
Sentiment scores and the number of geotweet in the selected ten states

We further examined the absolute value of the average positive and negative sentiment scores by states in Figure 4. In the majority of the states, the absolute value of the positive sentiment score is larger than that of the negative sentiment score. The difference between the positive and negative sentiment scores is relatively more obvious in the mainland states of Utah, Colorado, Nebraska, Minnesota, Connecticut, as well as in Hawaii and Alaska; the potential drivers triggering such differences across states may relate to information or news spreading locally or might be subject to the variations caused by the different sampling size in each state.

**Figure 4.**
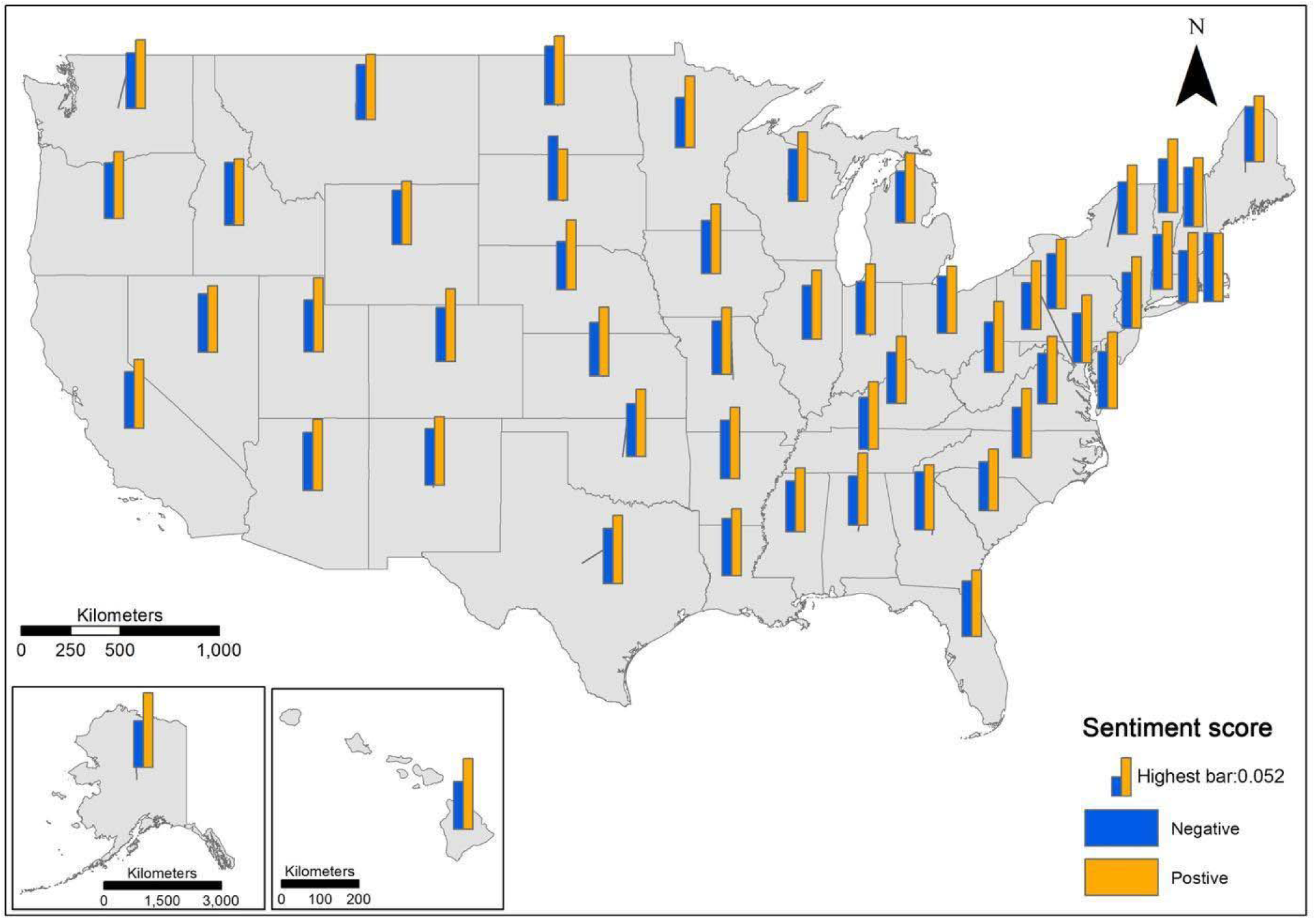
Negative and positive sentiment scores by state

The change in positive and negative sentiment scores over two periods of time (Phase 1 to Phase 2; Phase 2 to Phase 3) is compared and presented in Figure 5. From Phase 1 to Phase 2, the increase in positive sentiment scores (orange bars) appears in most states, most significantly in South Dakota, followed by North Dakota and Arkansas; meanwhile the decrease in negative sentiment scores (dark blue bars) is also observed in the majority of states, most significantly in South Dakota and Rhode Island, followed by Montana, North Dakota and Arkansas. From Phase 2 to Phase 3, the decrease in negative sentiment scores (light blue bars) appears in most states, most significantly in Idaho and Rhode Island, followed by North Dakota, Vermont, and New Hampshire. However, the change in positive sentiment scores (red bars) from Phase 2 to Phase 3 varies across states, with a slight increase that is more obviously observed in Idaho, North Dakota, New Mexico while a slight decrease is more obviously observed in South Dakota, Rhode Island, and Connecticut. In addition, the magnitude of both positive and negative sentiment scores from Phase 1 to Phase 2 (the height of dark blue and orange bars) is more obvious in most states than that of Phase 2 to Phase 3 (the height of light blue and red bars). It indicates that the fluctuation of people’s opinion on vaccines become less obvious with the gradual development of vaccines, accompanied by more encouraging news.

**Figure 5.**
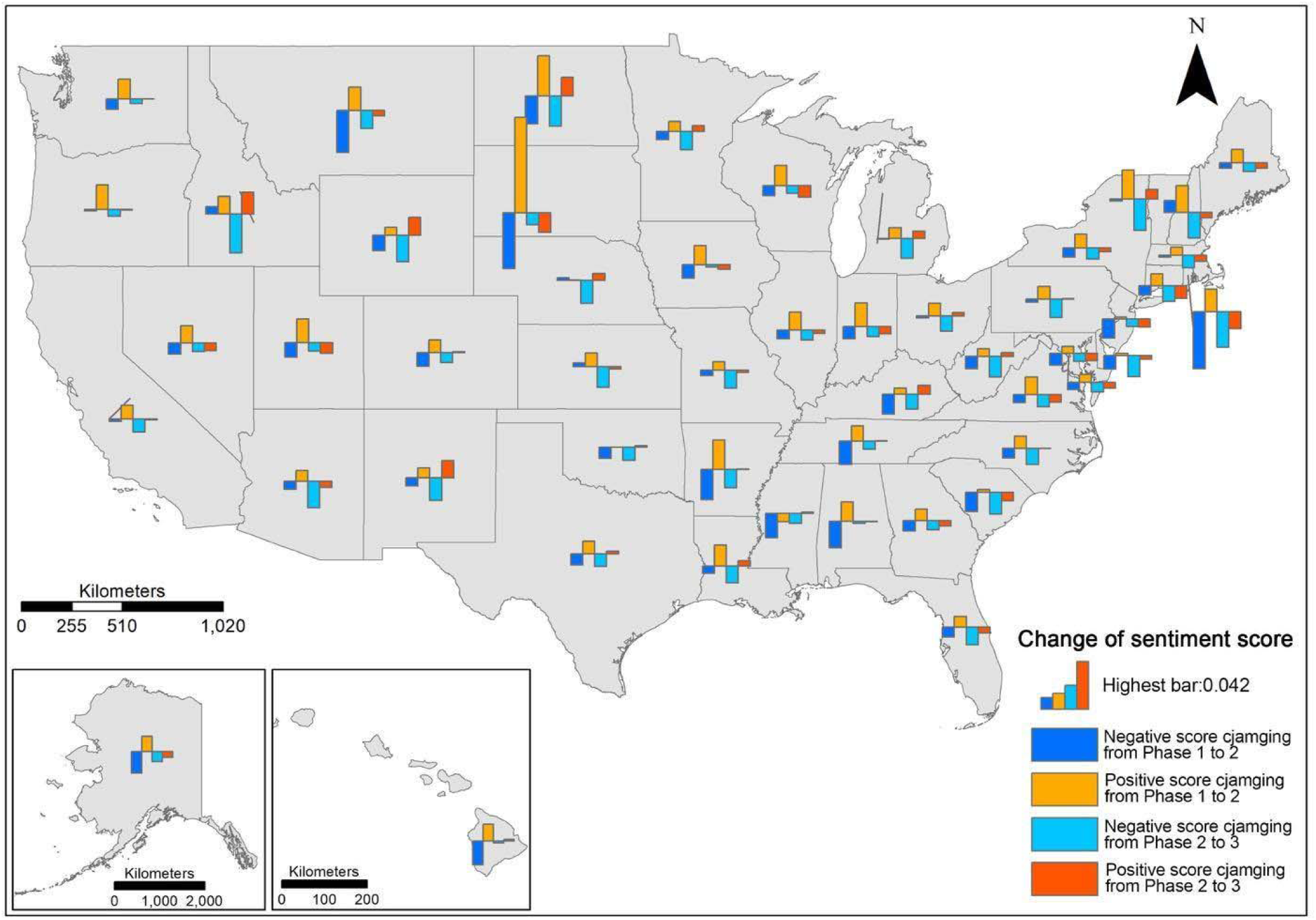
Change of sentiment over three phases

### 4.2 Emotion analysis

Figure 6 shows the temporal patterns of the eight types of emotion, including joy, trust, anticipation, trust, surprise, disgust, sadness, and fear. Through the vertical comparison of the weekly average trend lines (dash lines), we find that the emotion with the highest weekly average scores along the majority of the timeline is trust (blue dash line), followed by fear, anticipation, sadness, anger, joy, disgust, and surprise. It is worth noting that the weekly average emotion score of fear is higher than that of trust before the mid of April 2020, possibly due to the rapid infection and the ineffective control of viral spread at the early stage of the pandemic, which may cause fear, uncertainty or even feelings of panic [43]. Although the fluctuations of emotion scores (e.g., local peaks and valleys) can be found within each type of emotion, the overall tendency of the eight types of emotion implies the trustiness and anticipation of the public to vaccination, accompanied by the mixture of fear, sadness, and anger.

**Figure 6.**
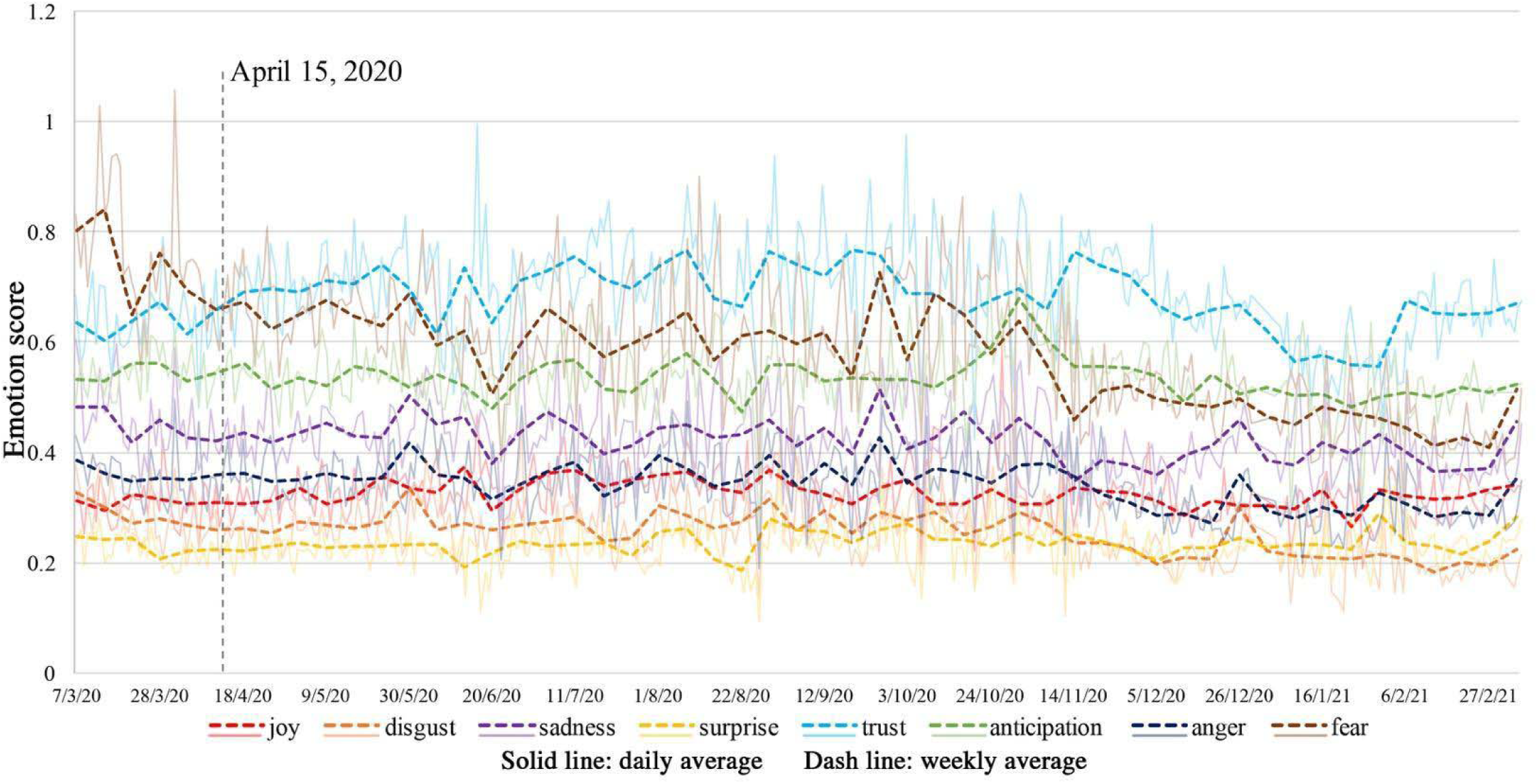
Daily and weekly average of emotion scores over the entire study timeline

We further investigate the relative distributions of eight emotions in each state, as indicated by the percentage of emotion scores for each type with different colors (Figure 7). The overall patterns of the eight emotions are consistent across most states—trust is the dominant emotion of the public responding to vaccination, followed by anticipation, fear, sadness, anger, joy, disgust, and surprise. The state-level patterns largely align with the national pattern revealed in Figure 6, although there are some exceptions with fear overweighting anticipation, joy and trust (e.g., Washington) and with fear, anger and sadness overweighing other emotions (e.g., Maine).

**Figure 7.**
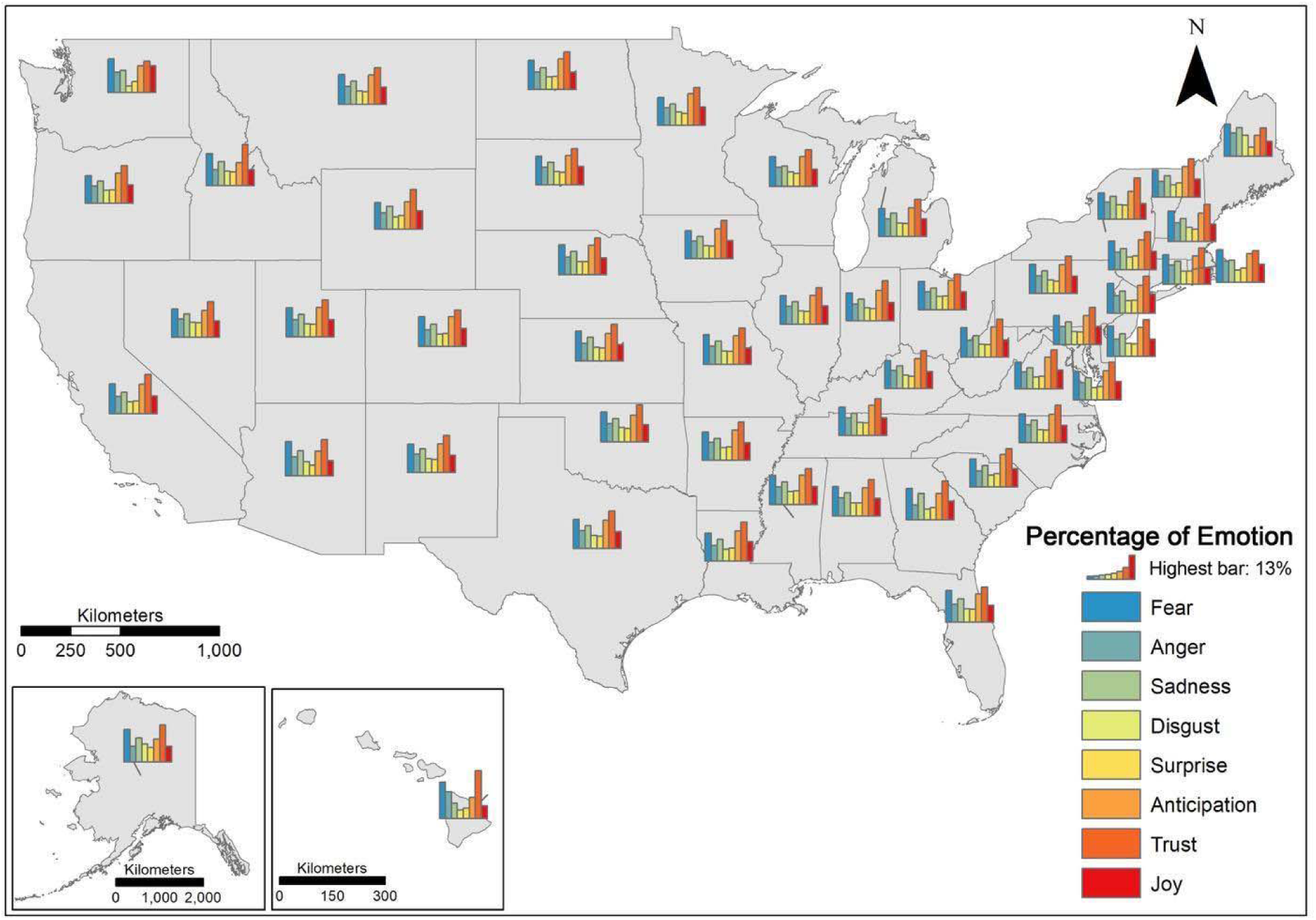
Eight types of emotion by state

We further compared the change in emotion percentage over two periods of time (Phase 1 to Phase 2; Phase 2 to Phase 3). From Phase 1 to Phase 2 (Figure 8A), the decrease in fear (dark blue bars) is observed in most states, though its magnitude varies across states: most significant in South Dakota, followed by North Dakota, Arkansas, Mississippi, North Carolina, and South Carolina. The change of anger, sadness, and disgust varies across states, decreasing in most states but increasing sporadically in some others (e.g., Idaho, New Mexico, and New Hampshire). Furthermore, with the decrease of trust, the increase in joy, trust, and anticipation is observed in most states except South Dakota. While it comes to the period from Phase 2 to Phase 3 (Figure 8B), it is difficult to generalize the pattern of emotion change across states in terms of its type and magnitude. The increase in joy, trust, anticipation and surprise along with the decrease in fear, anger, sadness and disgust are observed significantly (high bars) in Idaho and Rhode Island, followed by Missouri, Vermont, and New Hampshire. On the contrary, some states encounter a decrease in trust and anticipation in tandem with the increase in anger and sadness, including South Dakota, North Dakota, Montana, Kansas, Indiana, Maine, and Delaware. The complexity of emotion change from Phase 2 to Phase 3 varies across states, reflecting the diversity in people’s opinion and psychological reaction to vaccination, which would be subject to an in-depth investigation of causality.

**Figure 8(A):**
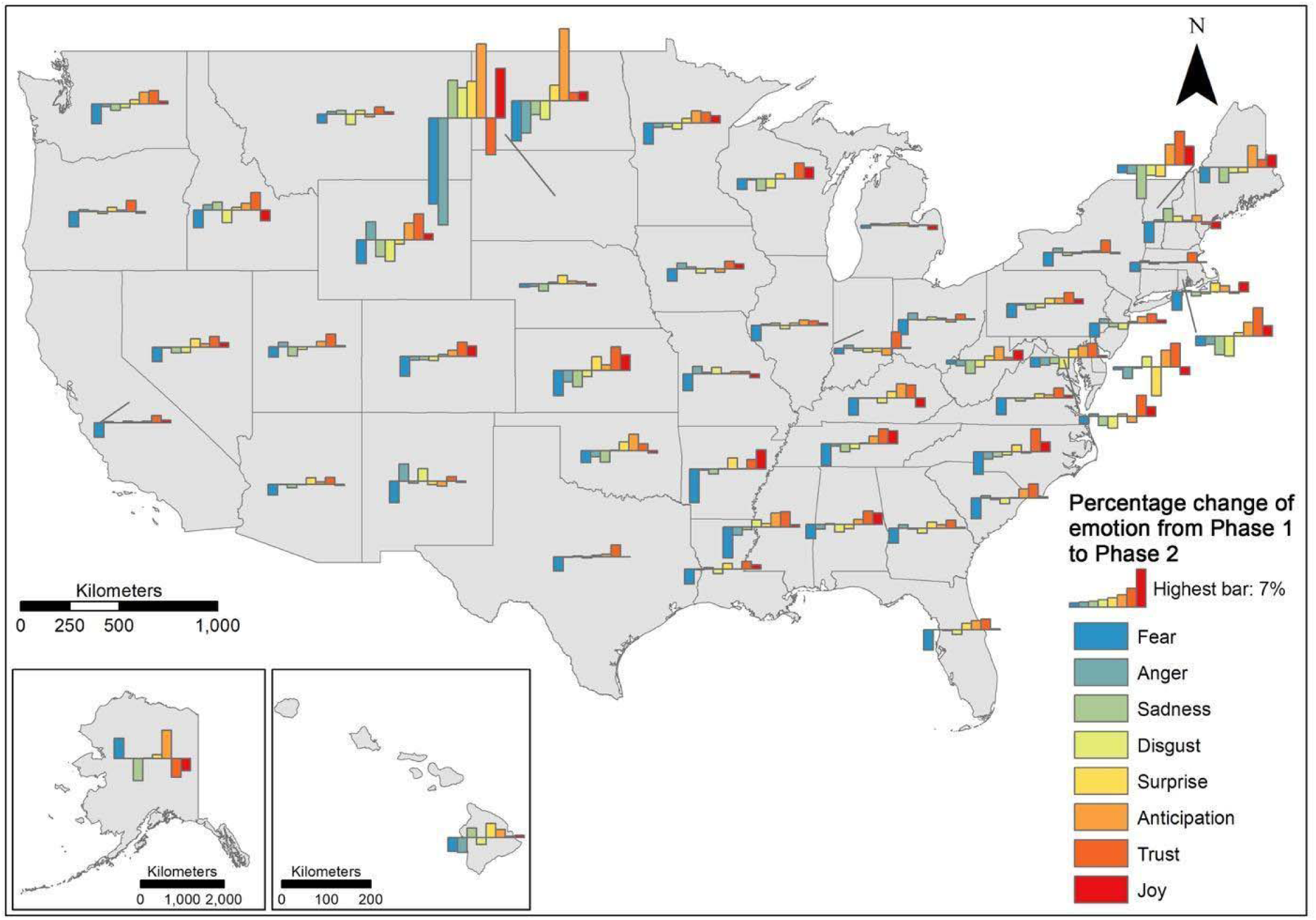
Change of emotion from Phase 1 to Phase 2

**Figure 8(B):**
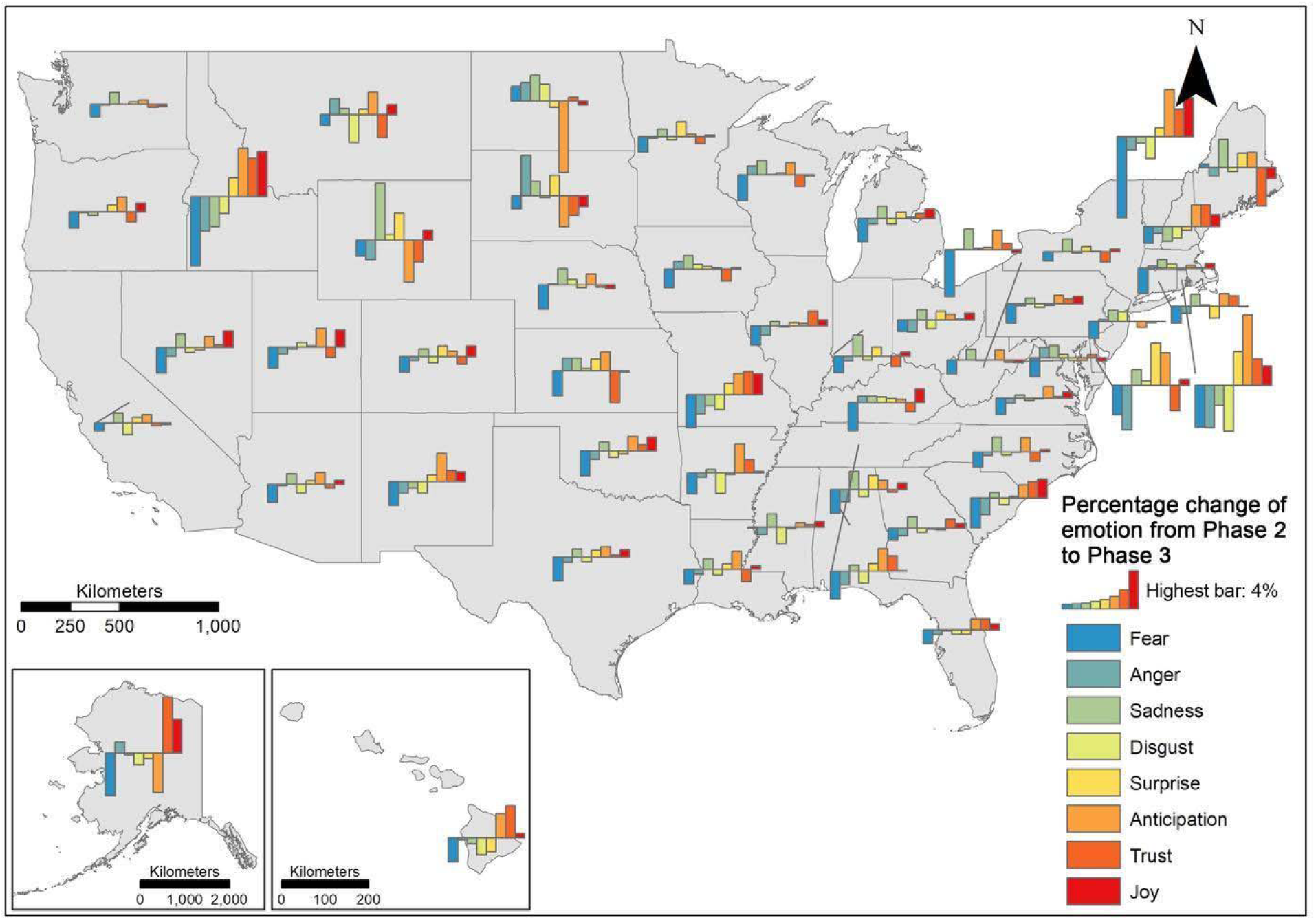
Change of emotion from Phase 2 to Phase 3

### 4.3 Word cloud visualization

We conducted word cloud mapping of the 50 popular words associated with positive and negative sentiment, respectively, over the three phases (Figure 9). The size of a word represents its popularity and the frequency with which the word appears in Tweets. Among the words associated with the positive sentiment, the popular ones are “hope”, “help”, “thank”, “love”, “safe”, “cure”, and “free”, although the word “peopl” with a more neutral nature, appears to be the largest one. Throughout the three phases, “hope”, “safe”, and “thank” grow larger from Phase 1 to Phase 3; in particular, “thank” becomes the most popular word in Phase 3. On the contrary, “flu”, “death”, “trump”, “fuck”, “lie”, “die”, “kill”, “shit”, and “stupid” are the popular words associated with the negative sentiment. Over the three phases, “flu” becomes smaller from Phase 1 to Phase 3 whereas “die”, “fuck”, “shit”, and “trump” evolve to be larger from Phase 1 to Phase 3; in particular, “trump” becomes predominant in Phase 2 possibly due to Trump’s increasing popularity, caused by the 2020 Presidential Election. More specifically, while people were waiting for the news of development in COVID-19 vaccines during Phase 1, their uncertainties on potential vaccines reflected on the included keywords, which target on the coronavirus and public’s frustration of the pandemic (e.g., “viru”, “death”, “cure” and “test”). Some keywords related to COVID-19 vaccine were also observed, including “hope” and “develop”. Positive news about the development of COVID-19 vaccines appeared in Phase 2, which brought hope as well as misinformation regarding the vaccines to the public. At this stage, more specific information of COVID-19 vaccines were discussed (e.g., “Pfizer”, “effect”, “risk”, “develop”, and “approve”) compared with Phase 1. With Pfizer and Moderna approved during Phase 3, the public’s attention moved from vaccine development towards vaccine distribution (“distribution”, “wait” and “free”), effectiveness (“safe” and “risk”), and priority (“teacher”). In all three phases, public figures (e.g., “Trump”, “Biden”, and “Bill Gates”) have contributed to hot topics with impacts on both positive and negative sentiment.

**Figure 9.**
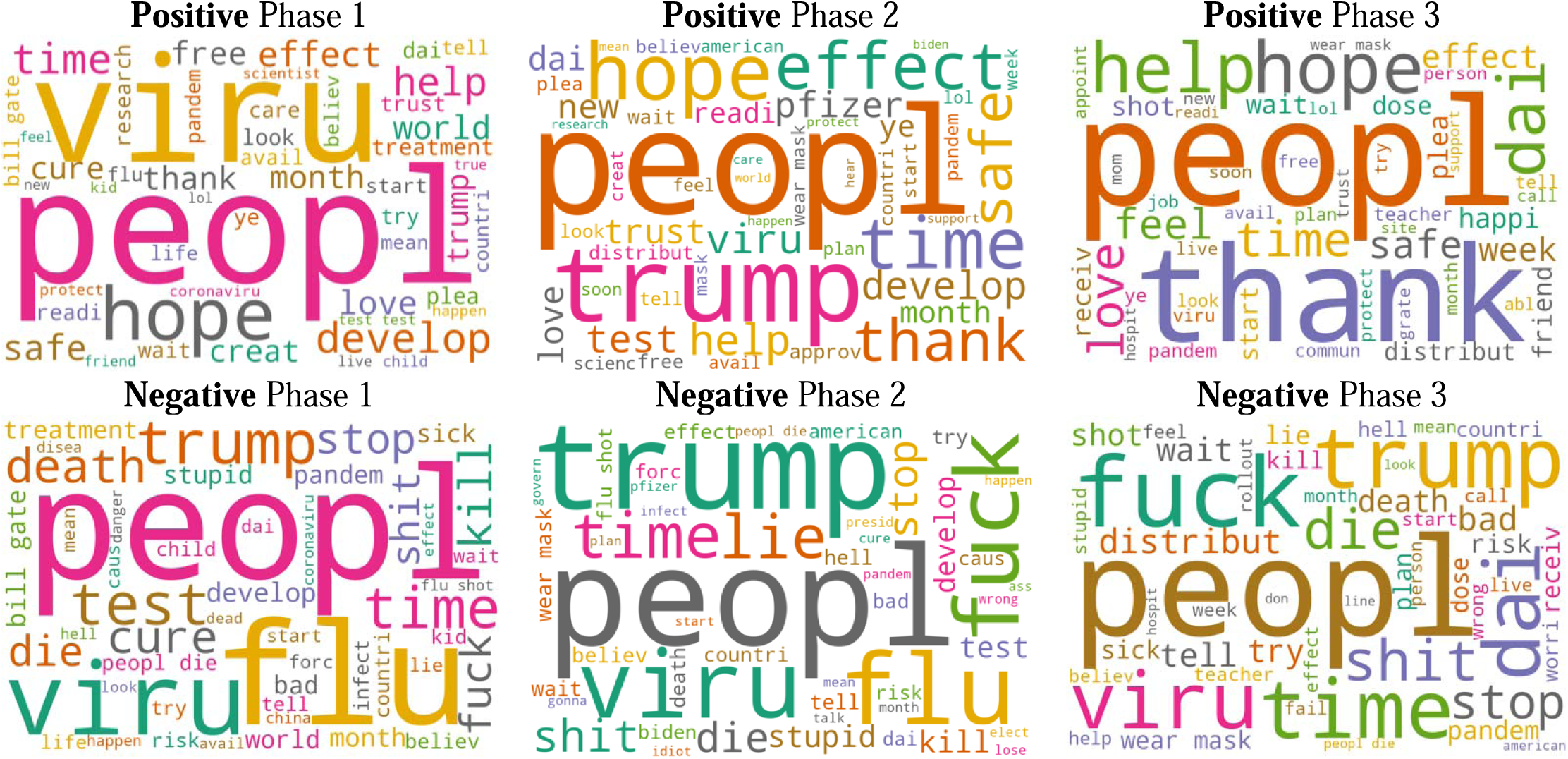
Popular keywords associated with positive and negative sentiment over three phases (Note: some keywords are extracted and presented as the main root of a word, e.g., “peopl” from people, “dai” from daily, and “viru” from virus)

## 5. Discussion and conclusion

### 5.1. Key findings

Drawing on geotweet from March 1, 2020, to February 28, 2021, this study aims to examine public opinion on COVID-19 vaccines in the US, through unveiling the spatiotemporal patterns of public sentiment and emotion over time, modeling the popular keywords and topics of Twitter contents, and analyzing the potential drivers of public opinion on vaccines. Our findings indicat that critical social/international events and/or the announcements of political leaders and authorities may have potential impacts on public opinion towards COVID-19 vaccines. In the proposed three phases over the study time, changes in public opinions on vaccines vary across space and time. The fluctuation in people’s sentimental response to the vaccine during the earlier stage of the pandemic is more obvious compared to that of the later stage in the pandemic, although the increase in positive sentiment in parallel with the decrease in negative sentiment are generally observed in most states, reflecting the raising confidence and anticipation of the public on vaccines. More specifically, the overall tendency of the eight types of emotion demonstrates the trustiness and anticipation of the public towards COVID-19 vaccines, accompanied by the mixture of fear, sadness and anger. The aforementioned findings reveal the diversity and complexity of people’s perception on and psychological reaction to COVID-19 vaccines, which further need great caution in the interpretation of analytical outcomes and the in-depth investigation of causality.

### 5.2. Implication and recommendation

The emergence of the Internet and social media has provided new platforms for persuasion and the rapid spread of (mis)information, which lead to new opportunities and challenges to the communication of vaccine information [44]. There are over 4.3 billion people using the internet nowadays, with 3.8 billion of these individuals as social media users [45]. The popularity of social media platforms coupled with the advent of digital detection strategies benefit public health authorities by enabling the monitoring of public sentiment towards vaccine relevant information in a geo-aware, (near) real time manner. This can inform more effective policy-making and promote participatory dialogue to establish confidence towards the vaccine, in order to maximize vaccine uptake. Some of our findings add new value to the current scholarship and also provide new insights and suggestions for policy implications with regard to safeguarding societal and economic health.

First, our findings indicate that critical social international events and/or the announcements of political leaders and authorities may have potential impacts on public opinion towards vaccines. Similarly, studies have also shown that vaccine-related content from popular figures is influential [46]. A study by Puri et al. (2020) [47] suggests that exposure to anti-vaccine content appears to have the most significant impact on downstream vaccine hesitancy amongst susceptible parents. We thus encourage and urge public figures to recognize the significance of sharing accurate and well-founded vaccine-related information on social media.

Second, our study reveals that vaccine-adverse conspiracy related to Bill Gate led to a significant decline in sentiment scores. We need to be aware of the fact that social media platforms with a massive number of users, to some degree, ‘disrupted’ traditional vaccine information communication [48], allowing anti-vaccination advocates to disseminate negative messages to a certain audience whose views on vaccination could be susceptible to change. However, it also means that governmental officials can take advantage of these platforms to communicate with individuals directly about the vaccination via geo-tailored messages to address the concerns specific to a certain region.

Third, while most states demonstrate similar trends in sentiment and emotion with a stable increase in positive attitudes towards COVID-19 vaccine, negative sentiments and emotions are more obvious in some states. Our geospatial analysis can help identify those areas, which need further research to address the underlying public fears and concerns with potential interventions. We further recommend the government, public health agencies, and relevant education programs to enhance the promotion of vaccines in such areas in order to provide positive guidance about vaccines as well as to minimize people’s concerns on vaccines.

### 5.3. Limitation and future work

Our study has several limitations that can be improved in future studies. First, the demographic of Twitter users is typically characterized by younger users who are avid users of mobile phone apps and the Internet, and such users may not be able to reflect the opinion and perception of the general public with varying demographics and socioeconomic status [49]. In addition, the representativeness of Twitter users is not stationary but geographically varying. Like other studies that rely on digital devices, the “Digital Divide” [50] issue needs to be acknowledged. This study only accounts for the reactions from Twitter users to vaccines, which neglect the underprivileged members of society to some degree, especially the poor, elderly, and those living in rural areas that do not have access to digital devices, as well as those who are not willing to share their thoughts on social media platforms. Additionally, the Twitter API we used allows limited access to approximately 1% of the total records. Future work needs to increase the sample size to reduce the uncertainties and fluctuations of sentiment scores and emotions. In early 2021, Twitter released a new Twitter API (academic research product track) that grants free access to full-archive search with enhanced features and functionality for researchers to obtain more precise, complete, and unbiased data for analyzing the public conversation [51]. Further efforts can be made to explore the potential of this new API in mining public opinions towards COVID-19 vaccines at finer-level scales.

Since emotion is a complex and integrated product of human feelings [52] future research efforts can be put into exploring more diverse dimensions of emotion in addition to the eight primary types of emotion. Moreover, disaster and crisis management include four phases, namely prevention (capacity building), preparation (early warning), response (search, rescue, and emergency relief) and recovery (rehabilitation) [50]. Management of the COVID-19 pandemic is still in the response phase. For policy and decision-making endeavors that are pertinent to COVID-19 crisis management, it will be highly beneficial if researchers and practitioners keep monitoring emotional and perspective variations throughout the response or even the recovery phase in the post-pandemic years. More importantly, to understand the impact of vaccination on countries, the workflow and methodology used in this study can be applied in multiple languages to global-scale geotweet.

## Data Availability

Users may contact the corresponding author to access the data.

## Abbreviations

API: application programming interface
GPS: global positioning system
LDA: Latent Dirichlet Allocation
NRCLex: National Research Council Canada Lexicon
SARS-CoV-2: severe acute respiratory syndrome coronavirus 2
VADER: valence aware dictionary for sentiment reasoning
WHO: World Health Organization

